# Hippocampal Synaptic Alterations Associated with Tau Pathology in Primary Age-Related Tauopathy

**DOI:** 10.1101/2023.02.22.23286323

**Authors:** Meaghan Morris, Gabrielle I Coste, Javier Redding-Ochoa, Haidan Guo, Austin R Graves, Juan C Troncoso, Richard L Huganir

## Abstract

Primary Age-Related Tauopathy (PART) is characterized by the aggregation of tau in the mesial temporal lobe in older individuals. High pathologic tau stage (Braak stage) or a high burden of hippocampal tau pathology have been associated with cognitive impairment in PART. However, the underlying mechanisms of cognitive impairment in PART are not well understood. Cognitive impairment in many neurodegenerative diseases correlates with synaptic loss, raising the question of whether synaptic loss occurs in PART. To address this, we investigated synaptic changes associated with tau Braak stage and a high tau pathology burden in PART using synaptophysin and phospho-tau immunofluorescence. We compared twelve cases of definite PART with six young controls and six Alzheimer’s disease cases. In this study, we identified loss of synaptophysin puncta and intensity in the CA2 region of the hippocampus in cases of PART with either a high stage (Braak IV) or a high burden of neuritic tau pathology. There was also loss of synaptophysin intensity in CA3 associated with a high stage or high burden of tau pathology. Loss of synaptophysin signal was present in AD, but the pattern was distinct from that seen in PART. These novel findings suggest the presence of synaptic loss in PART associated with either a high hippocampal tau burden or a Braak stage IV. These synaptic changes raise the possibility that synaptic loss in PART could contribute to cognitive impairment, though future studies including cognitive assessments are needed to address this question.

## Introduction

Primary age-related tauopathy (PART) is an age-associated neurodegenerative condition characterized by the aggregation of tau in the mesial temporal lobe [10]. Approximately 20% of individuals over 80 years of age have PART at autopsy. Cognitive impairment in PART is often mis-diagnosed as Alzheimer’s disease (AD) during life [21, 34]. The tau pathology in PART and AD are similar, with a similar molecular composition of the tau aggregates and a similar progression of tau pathology through the temporal lobe of the brain. However, unlike AD, the tau pathology in PART occurs in the absence of significant amyloid plaque deposition. Cognitive impairment occurs only in a subset of PART cases and has been associated with different measures of tau pathology. Some studies have shown an association between cognitive impairment and a high burden or density of tau aggregates in the hippocampus in PART [16, 22], while other studies have shown an association of cognitive impairment with a high tau pathology stage [2, 10, 18, 20].

The Braak system is used to stage tau pathology in PART and AD based on the progression of pathology through brain regions [5]. Classically, there are six Braak stages. Tau aggregates begin in the transentorhinal cortex (Braak I), then progress to involve the hippocampus and entorhinal cortex (Braak II), followed by involvement of the occipito-temporal gyrus (Braak III), and then the temporal cortex and other neocortical regions (Braak IV). The final two Braak stages (V and VI) involve the occipital cortex. While the highest two Braak stages are found in AD cases, these are rarely identified in PART [10]. PART cases with Braak stage IV tau aggregates, the highest stage typically observed in PART, have been associated with cognitive impairment [2, 4, 10, 18, 20, 32]. However, the underlying mechanisms of cognitive impairment in PART, particularly whether there is synaptic pathology or dysfunction, are largely unknown.

Synaptic loss is a common mechanism contributing to cognitive impairment in many human neurodegenerative diseases. In AD, synaptic loss correlates with both tau pathology [8, 23, 36] and cognitive decline [24, 28, 31, 33, 36]. In other neurodegenerative diseases with tau pathology, such as progressive supranuclear palsy and corticobasal degeneration, synaptic loss also correlates with clinical impairment [15, 30]. Studies in mouse models support the association of tau aggregation with synaptic loss [17]. However, the question of whether tau aggregation in PART leads to synaptic loss is largely unexplored.

To address the question of whether tau pathology is associated with synaptic alterations in PART, we compared synaptic immunolabeling in cases of definite PART with young controls and AD cases in the hippocampus. The tissue was immunolabeled for the pre-synaptic protein synaptophysin, which is widely used as a surrogate to measure synaptic loss in human tissue [28, 30, 33], and for phospho-tau (pS202/T205, AT8 antibody). As cognitive impairment in PART has been associated with both the burden of hippocampal tau pathology and Braak stage, we compared both measures to changes in the synaptophysin signal in multiple discrete regions of the hippocampus.

## Materials and Methods

### Human Tissue

All human tissue used in this study was from the Johns Hopkins Brain Resource Center (Baltimore, MD) with a post-mortem interval of less than 24 hours (Table 1). Control cases were defined as individuals under the age of 50 years with no evidence of amyloid deposition and no phosphorylated tau in the hippocampus (PHF1, gifted from Dr. Peter Davies). PART cases were selected to have hippocampal tau pathology (Braak stage II+) with no amyloid deposition (Thal 0, CERAD 0). AD cases were all selected for a high level of AD pathology (Braak V-VI, CERAD frequent) and were age-matched to the PART cases. Modified Bielschowsky silver staining [42] was used to determine Braak stage in PART and AD. Cases were excluded from all categories in this study if there was Lewy body disease, TDP-43 pathology, frontotemporal dementia, hippocampal age-related tau astrogliopathy, or hippocampal ischemic pathology.

**Table 1.** Demographic Information

| Diagnosis (n) | Braak | CERAD | Age | Sex (M:F) | PMI (hours) |
| --- | --- | --- | --- | --- | --- |
| Control (6) | 0<sup>a</sup> | 0 | 35.8 (27-43) | 4:2 | 17.5 (12-21) |
| AD (6) | V (n=2)<br>VI (n=4) | C | 85.7 (75-94) | 3:3 | 9.1 (3-17.5) |
| PART (12) | II (n=6) | 0 | 84.2 (69-91) | 3:3 | 13.9 (5-23) |
|  | III (n=3) | 0 | 79 (63-88) | 2:1 | 15 (12-16) |
|  | IV (n=3) | 0 | 86.7 (77-95) | 3:0 | 12.3 (10-14) |

AD, Alzheimer's disease; F, female; M, male; PART, Primary age-related tauopathy; PMI, post-mortem interval
<sup>a</sup>One control case is stage 1b by phosphorylated tau immunostaining using the updated Braak staging criteria [6].

### Antibodies

The following antibodies were used for immunofluorescence: synaptophysin (1:50, SY38, Abcam, Cambridge, UK), NeuN (1:500, ABN78, Millipore Sigma, Burlington, MA), AT8 (pS202/T205) phospho-tau (MN1020, Thermo Fisher Scientific, Waltham, MA). Prior to use, the AT8 antibody was concentrated using a centrifugal filter (UFC9010, Millipore Sigma, Burlington, MA) then covalently labeled with Alexa 594 using an antibody labeling kit (A20185, Thermo Fisher Scientific, Waltham, MA) according to the manufacturer’s instructions. Labeled antibody was used at a dilution of 1:800.

### Immunofluorescence

Formalin-fixed, paraffin-embedded tissue sections were cut at 5 µm from the mid-hippocampus at the level of the lateral geniculate nucleus. Tissue sections were deparaffinized and citrate antigen retrieval (ImmunoRetriever with citrate, Bio SB Inc, Goleta, CA) was performed at high heat and high pressure for 15 minutes (TintoRetriever Pressure Cooker, Bio SB Inc, Goleta, CA). Sections were labeled first with antibodies against synaptophysin and NeuN, then with secondary antibodies Alexa 488 goat anti-mouse IgG1 (1:400, A21121, Invitrogen Thermo Fisher Scientific, Waltham, MA) and Alexa 647 goat anti-rabbit (1:400, A21245, Invitrogen Thermo Fisher Scientific, Waltham, MA), followed by a 1-hour incubation with the labeled AT8 antibody. Autofluorescence was quenched using TrueBlack Plus Lipofuscin Autofluorescence Quencher (Biotium, Fremont, CA) diluted 1:40 in 70% ethanol and applied for 50 seconds. Hippocampal regions were marked based on anatomic landmarks by a Board-certified neuropathologist. Images were taken at 400X on a Zeiss LSM 880 confocal microscope (Zeiss, Oberkochen, Germany), with 1-3 images taken per region per case in the molecular layer of the dentate gyrus, stratum radiatum of CA1 and CA2, and the stratum radiatum/lucidum of CA3. Image acquisition parameters were uniform across the entire cohort. Experimenters were blinded during imaging and processing.

### Imaris Analysis

Images were analyzed using Imaris version 9.8.2 (Oxford Instruments, Abingdon, UK), with a region of interest (ROI) selected to exclude neuronal cell bodies (NeuN+) and large vessels. Two ROIs were generated in the molecular layer: the outer molecular layer was included in the analysis of the dentate gyrus synaptophysin intensity while the inner molecular layer was used for synaptophysin intensity normalization within each case. Synaptophysin intensity was calculated using the synaptophysin channel “intensity mean” parameter for each ROI, which was normalized to the average “intensity mean” of the inner molecular layer in that case. Tau pathology was calculated as a percent volume within the ROI, with neuritic tau pathology volume measured in each region using the surface function in Imaris (Online Resource 1: Supp. Table 1, Supp. Fig. 1A). The tau score was calculated as the sum of average tau pathology per region per case. High tau in PART was defined as a tau score equal or above the minimum tau score of the AD cases (score 0.73). All other PART cases were defined as low tau.

### XTC Image Restoration and Analysis

To accurately detect fluorescent signals from dense networks of thousands of individual synaptic puncta, images were denoised using a previously trained image restoration algorithm optimized for signal from fluorescently labeled synaptic proteins (Online Resource 1: Supp. Fig. 1B) [41]. Briefly, this deep learning algorithm based on Content Aware Image Restoration (CARE) [39] was trained to discern properties of fluorescently labeled synaptic proteins using paired low- and high-resolution images of the same live tissue. This cross-modal trained CARE (XTC) algorithm restores optimal, super-resolution-like image quality from sub-optimal resolution images [41]. By running the synaptophysin images through this algorithm, the signal-to-noise ratio was improved to match Airyscan-like resolution, allowing ilastik to segment individual puncta with high accuracy using the density counting workflow (https://www.ilastik.org/documentation/counting/counting)[3].

### Statistical Analysis

Data were analyzed in R (version 4.2.0) / RStudio (Posit, Boston, MA), using nlme (version 3.1-159) and lme4 (version 1.1-30) [1, 26]. The data were analyzed with mixed effects linear modeling with “∼1|Case” to model random effects. For analysis of the effects of Braak stage and neuritic tau burden, each mixed effects linear model included an interaction term for hippocampal region, with sex as a co-variate. Age was only included as a co-variate in the Braak stage analysis, as it showed a linear correlation with neuritic tau burden. For AD cases, each mixed effects linear model included an interaction term for hippocampal region and diagnosis, with sex as a co-variate. GraphPad Prism 9.4.1 (GraphPad Software, San Diego, CA) was used for all graphs, and for simple linear regression and ANOVA analyses. All points on the graphs represent the average value for that region for a single case.

## Results

To determine whether tau pathology in PART is associated with synaptic alterations in the hippocampus, we quantified neuritic tau pathology and synaptophysin in PART and compared them to young control cases and AD cases (Fig. 1A). Only cases of definite PART were used with no evidence of amyloid deposition (CERAD score 0, Thal score 0) (Table 1) [10]. Synaptophysin puncta were quantified after cross-modality image restoration (XTC) [41] (Supp. Fig. 1B). The synaptophysin fluorescence signal was normalized to the average signal from the inner molecular layer of the dentate gyrus, according to previous literature [28]. Images were taken exclusively from the stratum radiatum of the CA1-2 regions, the stratum radiatum/lucidum of CA3, and the molecular layer of the dentate gyrus. These regions were selected because they included several regions previously shown to have synaptic changes in AD [27, 28, 36], with the addition of the CA2 region, which is preferentially affected in PART [38]. As the pyramidal region was not imaged, the burden of tau pathology in each case was determined based on the percent volume of neuritic phospho-tau.

**Figure 1.**
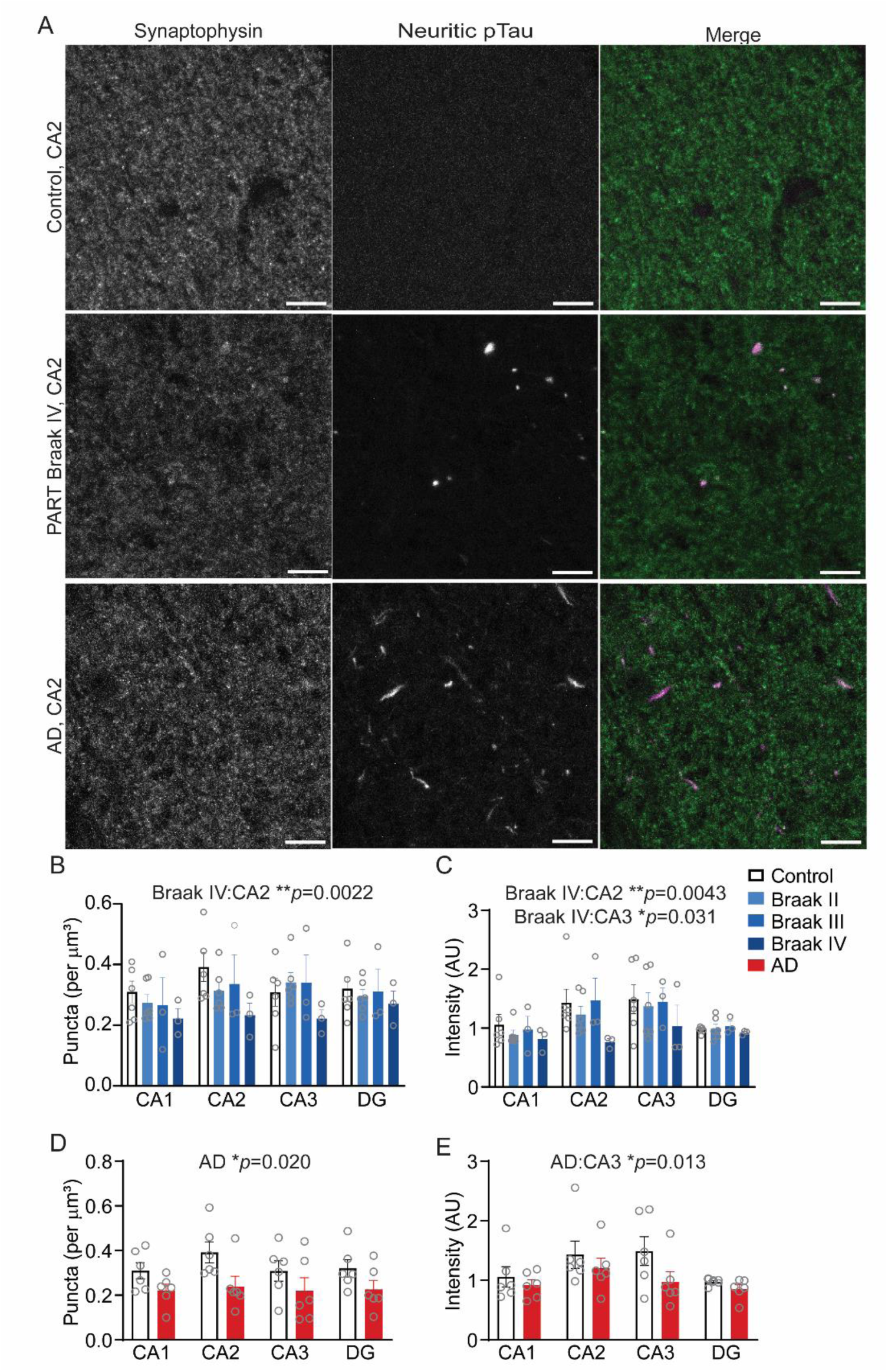
Loss of Synaptophysin in CA2 with Braak Stage IV in PART. (A) Immunostaining for synaptophysin and phospho-tau in CA2 of the hippocampus in a control, Braak IV PART, and high-level (Braak VI) AD. Quantification of synaptophysin puncta (B) and intensity (C) by Braak stage in PART. Quantification of synaptophysin puncta (D) and intensity (E) in Alzheimer’s disease. Scale bars are 10 µm. Acquisition and display settings are standard across all images. Graphs show average value per region per case, with statistics shown from mixed effects linear model. AD, Alzheimer’s disease; DG, dentate gyrus; pTau, phosphorylated tau.

In control cases, the stratum radiatum of CA2 showed a greater number of synaptophysin puncta and higher intensity compared to other hippocampal subfields in all analyses (Table 2, Fig. 1B-C), which has been described previously in mice [29]. The stratum radiatum in CA3 showed a higher synaptophysin intensity, but not an increased number of synaptophysin puncta in control individuals (Table 2, Fig. 1B-C). There was also an effect of sex on synaptophysin puncta but not on synaptophysin intensity (Table 2), though the significance of this finding is unclear given the slight bias toward male controls and PART cases in this cohort. There was no association between post-mortem interval and synaptophysin puncta or intensity (Online Resource 1: Supp. Fig. 2).

**Table 2.** Effects of Tau and Disease on Synaptophysin in PART and AD

| Disease | Synaptophysin Measure | Disease Measure | Disease Effect | Interaction (Disease Measure*Region) |  |  | Region |  |  | Sex (M) | Age |
| --- | --- | --- | --- | --- | --- | --- | --- | --- | --- | --- | --- |
|  |  |  |  | CA1 | CA2 | CA3 | CA1 | CA2 | CA3 |  |  |
| PART | Puncta (per $\mu\text{m}^3$) | Braak Stage (II,III,IV) | II: 0.15<br>III: 0.16<br>IV: 0.18 | II: -0.0014<br>III: -0.026<br>IV: -0.029 | II: -0.053<br>III: -0.047<br>**IV: -0.11\*\*** | II: 0.055<br>III: 0.035<br>IV: -0.040 | -0.020 | **0.071\*\*\*** | -0.0090 | **-0.088\*** | -0.0040 |
| PART | Intensity (AU) | Braak Stage (II,III,IV) | II: 0.62<br>III: 0.60<br>IV: 0.59 | II: -0.14<br>III: -0.098<br>IV: -0.14 | II: -0.22<br>III: -0.020<br>**IV: -0.62\*\*** | II: -0.20<br>III: -0.17<br>**IV: -0.47\*** | 0.033 | **0.46\*\*\*** | **0.58\*\*\*** | -0.029 | -0.012 |
| PART | Puncta (per $\mu\text{m}^3$) | Tau Burden (L,H) | L: -0.012<br>H: -0.049 | L: -0.010<br>H: -0.019 | L: -0.048<br>**H: -0.090 \*\*** | L: 0.048<br>H: -0.0039 | -0.020 | **0.071\*\*\*** | -0.0093 | -<br>**0.099\*\*** | - |
| PART | Intensity (AU) | Tau Burden (L,H) | L: 0.062<br>H: -0.054 | L: -0.19<br>H: -0.042 | L: -0.15<br>**H: -0.44\*** | L: -0.14<br>**H: -0.42\*** | 0.031 | **0.46\*\*\*** | **0.58\*\*\*** | -0.094 | - |
| AD | Puncta (per $\mu\text{m}^3$) | Disease (AD) | **-0.12\*** | 0.015 | -0.059 | 0.00024 | -0.020 | **0.071\*\*** | -0.0094 | **-0.13\*\*** | - |
| AD | Intensity (AU) | Disease (AD) | -0.12 | 0.037 | -0.11 | **-0.46\*** | 0.031 | **0.46\*\*\*** | **0.58\*\*\*** | -0.019 | - |

Data are fixed effects estimates from the mixed effects linear models. AD, Alzheimer's disease; AU, arbitrary units; H, high tau neuritic burden; L, low neuritic tau burden; M, male; PART, primary age-related tauopathy.

**Figure 2.**
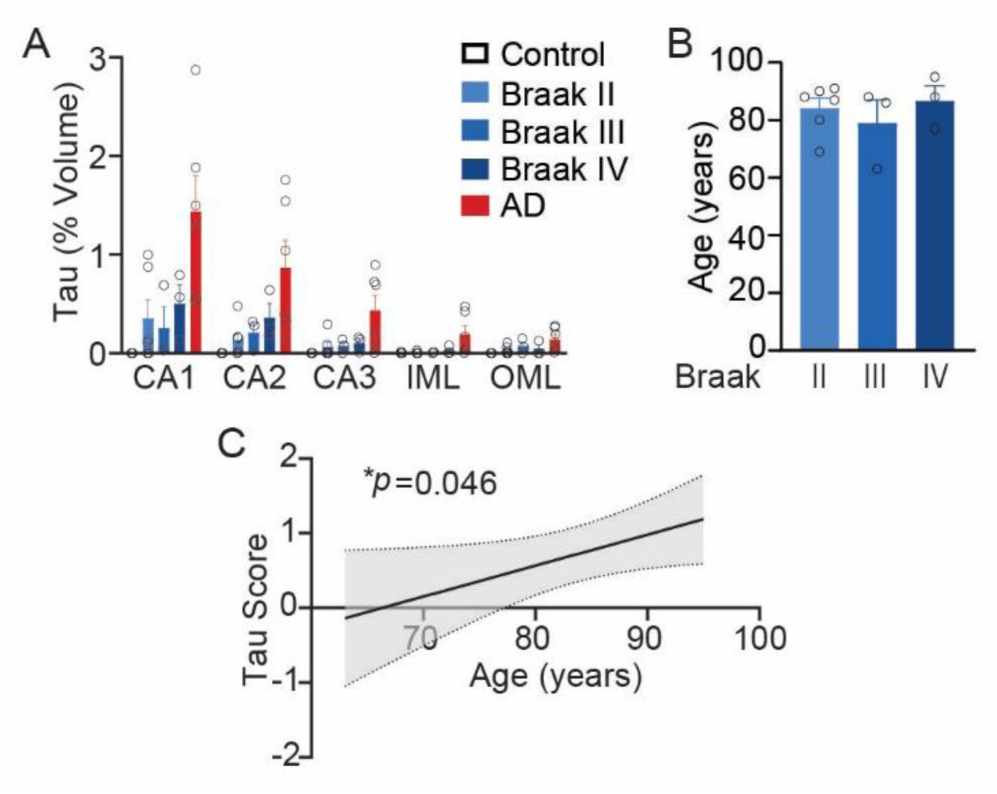
Neuritic Tau Pathology by Braak Stage and with Aging A) Quantification of phosphorylated tau volume by region and disease. (B) Age was not associated with Braak score in PART (*p*=0.64, one-way ANOVA). (C) Association of neuritic tau score with age in PART (*p*=0.046, simple linear regression, shade is 95% confidence interval). AD, Alzheimer’s disease; IML, inner molecular layer of the dentate gyrus; OML, outer molecular layer of the dentate gyrus.

There was loss of both the puncta number and synaptophysin intensity in the CA2 region with Braak IV PART relative to young control images from each region. Loss in CA3 was only detected by signal intensity but not in puncta number (Fig. 1B-C, Table 2). Braak stages II or III in PART did not show significant loss of synaptophysin puncta or intensity in any region. Age was not associated with synaptophysin loss in linear models which included Braak stage (Table 2). In AD, by contrast, there was significant loss of synaptophysin puncta across the hippocampus, though loss of synaptophysin intensity was only significant in CA3 (Fig. 1D-E, Table 2).

Neuritic tau pathology was higher in CA1 and CA2 in PART and AD compared to other hippocampal subregions (Fig. 2A). PART cases had a lower burden of tau pathology compared to AD cases, consistent with prior reports [38]. However, the neuritic tau burden varied considerably within each Braak stage for PART, such that increasing Braak stage was not associated with increased neuritic tau burden. Though age did not vary across Braak stages in this cohort (Fig. 2B), there was a linear association between increasing age and the burden of neuritic tau pathology (Fig. 2C). Due to the correlation between age and neuritic tau, age was not included as a covariate in subsequent analyses with tau burden.

To assess the effect of a high burden of tau pathology on synaptophysin in the hippocampus, the PART cases were divided into high tau burden (H Tau) and low tau burden (L Tau). A score for neuritic tau pathology was created in each case by adding the average tau volume in each hippocampal region. PART cases with a tau score at least that of the lowest AD case were classified as high tau burden, and the remaining cases were classified as low tau burden (Fig. 3A). Using this classification, 7 cases were classified as low tau burden and 5 cases were classified as high tau burden. Aligning with the prior observation that Braak stage and neuritic tau burden were not strictly related, both groups were comprised of a mix of Braak stages. The low tau burden group included 4 Braak II, 2 Braak III, and 1 Braak IV cases, while the high burden tau group included 2 Braak II, 1 Braak III, and 2 Braak IV cases.

**Figure 3.**
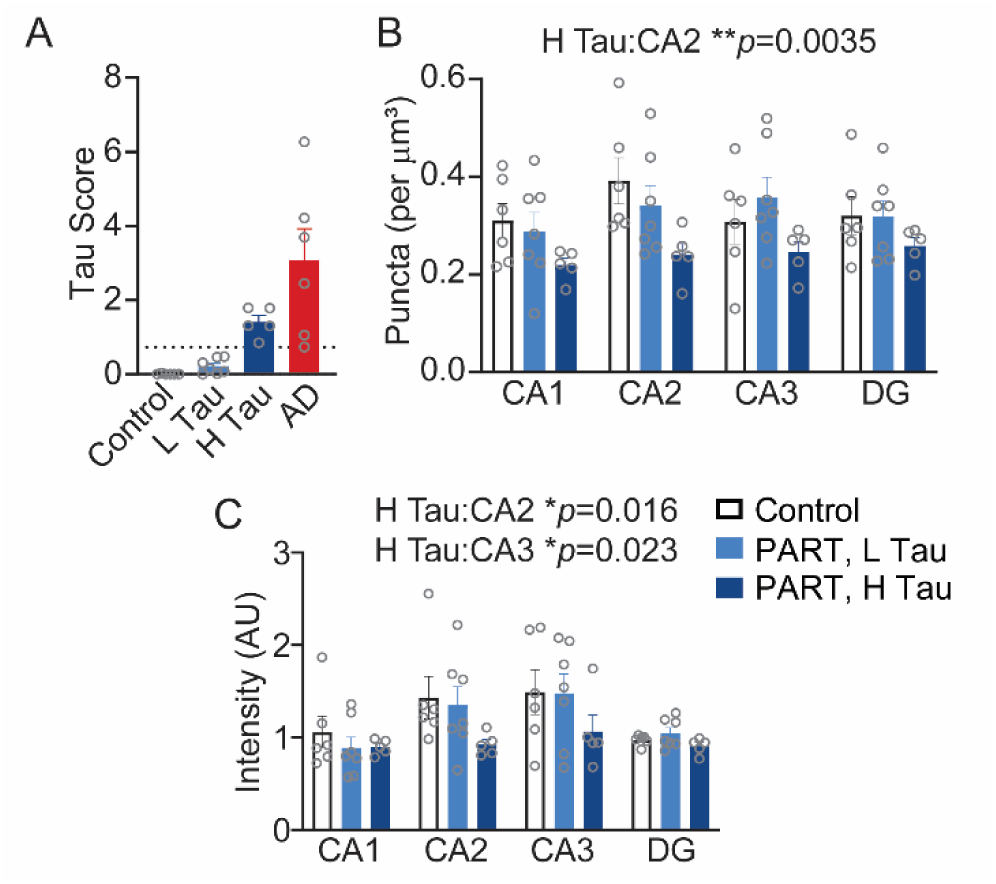
Loss of Synaptophysin with High Neuritic Tau Burden in PART. (A) Hippocampal neuritic tau score used to divide PART cases into high and low tau burden (line is threshold between high and low tau scores). Quantification of synaptophysin puncta (B) and intensity (C) by tau burden in PART. Graphs show average value per region per case, with statistics shown from mixed effects linear model. AD, Alzheimer’s disease; DG, dentate gyrus; H Tau, high neuritic tau score; L Tau, low neuritic tau score; pTau, phosphorylated tau.

Analysis of the synaptophysin signal by tau burden in PART also showed effects selectively in CA2 and CA3. PART cases with a high tau burden showed loss of both synaptophysin puncta and intensity in CA2, with loss of synaptophysin intensity in CA3 (Table 2, Fig. 3B-C). There was no effect on synaptophysin puncta or intensity in PART cases with a low hippocampal neuritic tau burden.

## Discussion

Primary age-related tauopathy (PART) has been associated with cognitive impairment, but the underlying mechanism is not well understood. In particular, synaptic alterations which might contribute to cognitive impairment in PART are unknown. In this study, we identified loss of synaptophysin puncta and intensity in the CA2 region of the hippocampus in cases of PART with either a high stage (Braak IV) or a high burden of neuritic tau pathology. There was also loss of synaptophysin intensity in CA3 associated with a high stage or high burden of tau pathology. These novel findings suggest the presence of synaptic loss in PART which correlates with either a high hippocampal tau burden or a Braak stage IV. These synaptic changes raise the possibility that cognitive impairment in PART may be associated with synaptic loss.

This study presents evidence for synaptic loss in PART associated with tau pathology. While there was also synaptic loss in AD in this study, in agreement with prior literature [27, 28, 31, 33], the pattern of synaptic changes in PART was distinct. The synaptic effects of AD were more global, while PART specifically affected synapses in CA2 with a possible smaller effect in CA3. The synaptic alterations in PART were also only associated with either a Braak IV stage or a high burden of neuritic tau pathology. Despite the presence of hippocampal tau pathology in all PART cases (Braak II+), cases with a lower tau stage or lower neuritic tau burden did not show significant loss of synapses in any quantified region. There was also no association of synaptophysin signal with age if Braak stage was accounted for, consistent with previous literature [14]. Synaptic changes were therefore specific to individuals with either a high Braak stage (Braak IV) or a high burden of hippocampal neuritic tau pathology.

Unfortunately, cognitive data was not available for many of the cases in this study. This limits our ability to assess whether synaptic loss in PART was associated with cognitive decline directly. The same factors associated with evidence of synaptic loss in this study, namely Braak IV stage and a high burden of hippocampal tau pathology, are also associated with cognitive impairment in PART [10, 16, 18, 20, 22]. However, there were only a small number of cases in this study and few of them had cognitive data. Studies with more cognitive data are needed to determine if there is a direct association between synaptic loss in the hippocampus and cognitive impairment in PART.

The loss of synaptophysin specifically in CA2 in PART is notable given that multiple studies have suggested CA2 is selectively vulnerable to tau pathology in PART [19, 37, 38]. This proclivity for CA2 is shared by other non-AD neurodegenerative diseases [25, 38], though the biologic basis is not well understood. CA2 is a unique region in the hippocampus about which relatively little is known. Prior studies in animal models show that neurons in CA2 have distinct molecular properties and patterns of gene expression compared to other hippocampal regions [25]. In humans, CA2 is more resistant to hypoxic-ischemic, traumatic, and excitotoxic injury compared to other hippocampal regions [13]. In this study, we also provide evidence that stratum radiatum of CA2 may have a greater synaptic density than other hippocampal regions. While not well studied in humans, this higher CA2 synaptic density has been reported in mice [29].

The effects of synaptic loss in CA2 and the accumulation of tau pathology in this region in PART are unknown. Lewy body disease, which selectively accumulates synuclein in the CA2 region in the hippocampus, has shown an association between CA2 pathology and dementia [7, 25]. Rodent models support a role for CA2 in cognition, though it appears to be less critical for traditional hippocampal-based memory tasks. Though activated in classic hippocampal memory tasks in rodents, lesions of CA2 do not impair performance on those tasks [13], making the precise role of CA2 difficult to define in hippocampal memory. However, similar studies show a critical role of CA2 in social recognition memory [13]. Facial recognition tasks represent a potentially similar test in humans. Intriguingly, facial recognition is known to be impaired in Lewy body disease patients [9] and in aging [11]. As facial recognition tasks are not tested regularly in aging cohorts, the effects of tau pathology in PART on this task are unknown, though this potentially represents a new avenue of investigation for tau-associated cognitive impairments in PART.

The changes in synaptophysin labeling, a pre-synaptic marker, may reflect pathology in the afferent connections in this region. Despite a similar burden of neuritic tau pathology in CA2 and CA1, significant synaptic changes were not identified in CA1. CA3 showed a lower burden of neuritic tau pathology than CA1, but did have loss of synaptophysin intensity. This implies that the synaptic findings in this study may reflect pathology affecting the afferent connections to CA2 and CA3 rather a local effect of neuritic tau. Layer II neurons in the entorhinal cortex and the locus ceruleus both innervate CA2 and CA3 [12, 13, 35] and are regions which show tau pathology early in PART [10]. Both CA2 and CA3 are also innervated by the dentate gyrus and innervate each other, though the burden of neuritic tau pathology in the dentate and CA3 is lower and less likely to account for the synaptic changes. They are both innervated by inputs from the septal nuclei [12, 40], which are less studied in PART pathology, as are the other afferent connections to CA2, including hypothalamic and supramammillary nuclei [12]. Further studies are needed to determine which synaptic connections are specifically affected in PART and whether these alterations are associated with tau pathology in the afferent neuronal population.

Though Braak stage IV and a high burden of neuritic tau pathology in the hippocampus were both associated with synaptic changes in this study, Braak stage and the burden of neuritic tau pathology were not directly related. There was a large variability in the burden of hippocampal neuritic tau pathology among cases with the same Braak stage. Braak staging reflects the brain regions affected by tau pathology, specifically neurofibrillary tangles, but does not incorporate density of tau pathology in its staging system. The neuritic tau burden increased with age, consistent with prior studies [16, 22, 38]. Unlike some prior studies [10, 18], there was no association between Braak stage and age in this small cohort. While most studies have found an association between Braak stage and cognitive impairment [4, 10, 18, 32], some studies found only an association with hippocampal tau burden but no association with Braak stage [16, 22]. Further studies are needed which include both measures of tau pathology to clarify if one of the two is more reliably associated with cognitive impairment and synaptic loss in PART. If hippocampal tau burden were more reliably associated with synaptic loss and cognitive impairment, it would suggest that measures other than Braak stage should be used to assess disease severity in PART.

There were some regional differences between the measurements produced by synaptophysin puncta and synaptophysin intensity. This could partly be due to biologic factors. A subset of the synapses in CA3, specifically the mossy fiber synapses, are known for the unusually large size of the pre-synaptic boutons. So an increased synaptophysin intensity without an increase in puncta in CA3 in controls in this study compared to other hippocampal regions may relate to an increased size of some CA3 synapses. Another difference in the two synaptophysin measurements could be the normalization of the intensity within each case. While this minimized variability between cases, normalization can also alter the magnitude of changes in any individual regions. For instance, in AD, there was a trend toward lower synaptophysin intensity in all regions which was significant in CA3, but the puncta measurements showed global loss of synaptophysin puncta in the hippocampus.

In summary, this study provides novel evidence of synaptic loss in PART associated with a high burden of neuritic tau pathology or Braak stage IV disease. There was no evidence of synaptic changes in cases with a lower stage or burden of hippocampal neuritic tau pathology. This synaptic loss specifically affected the hippocampal CA2 region, with some evidence of synaptic alterations affecting CA3. The vulnerability of CA2 to synaptic alterations correlates with previous studies noting CA2 is selectively vulnerable to tau pathology in PART. This study also raises the possibility that synaptic loss in PART may contribute to cognitive impairment.

## Data Availability

All data is available upon reasonable request to the corresponding author.

## Acknowledgements

We would like to acknowledge the Multiphoton Imaging Core in the Johns Hopkins Department of Neuroscience for technical assistance and providing software resources for this study, and Jamie Huynh, Johns Hopkins University, for assistance with image analysis.

## Declarations

### Ethics Approval

The work in this study was approved by the Johns Hopkins Institutional Review Board and was performed in accordance with the 1964 Declaration of Helsinki and its later amendments.

### Data Availability

All data is available upon reasonable request to the corresponding author.

### Author Contributions

MM, JCT, and RLH designed the study. JRO, HG, GIC, and ARG contributed to methodology and data collection. MM, GIC, and ARG contributed to data analysis. MM wrote the manuscript. MM, GIC, JRO, HG, ARG, JCT, and RLH reviewed with editing. All authors have read and approved the final manuscript.

