## Supplemental Files for "Hippocampal Synaptic Alterations Associated with Tau Pathology in Primary Age-Related Tauopathy"

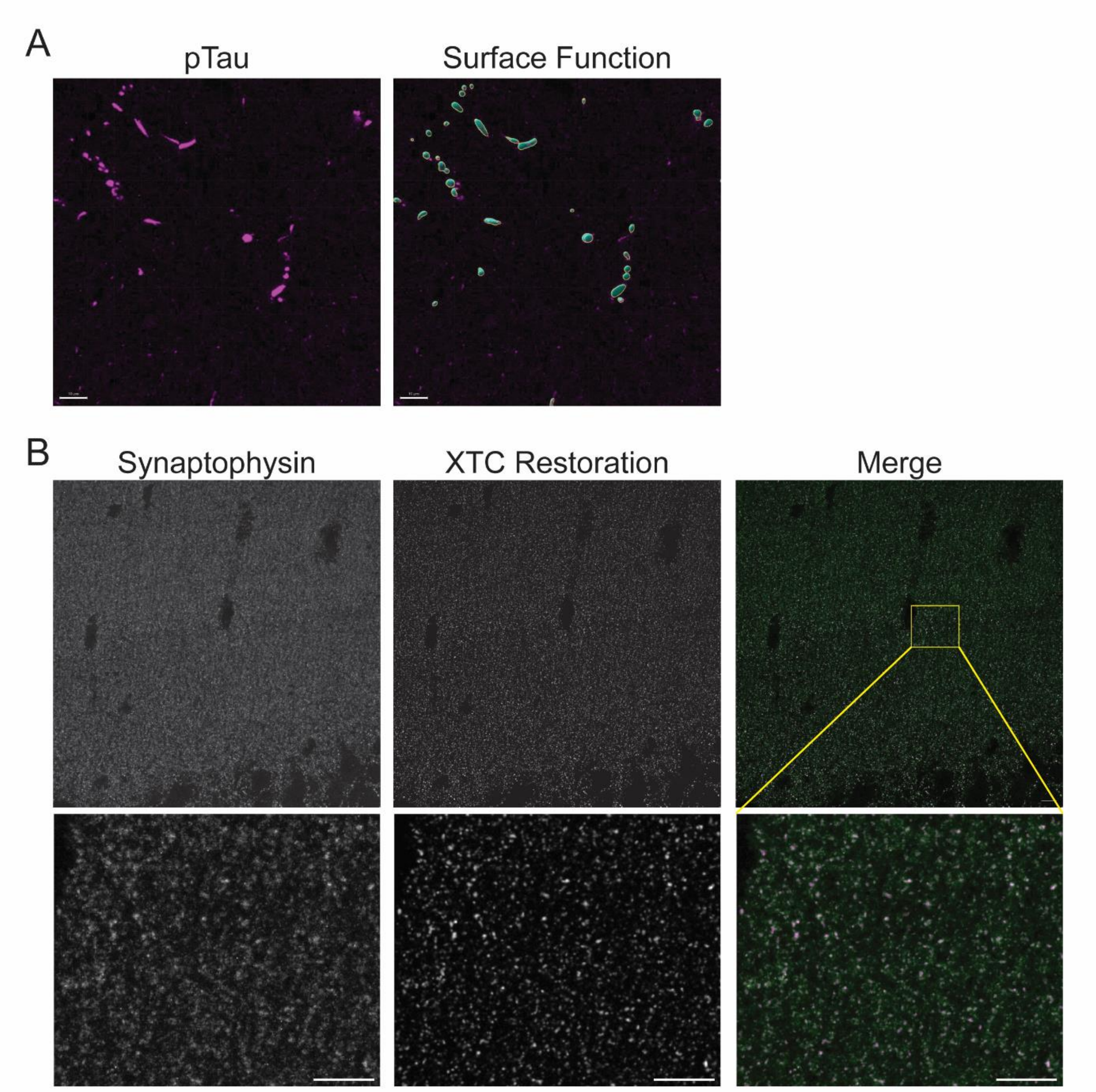


**Supplemental Figure 1**. Phosphorylated Tau Analysis and Synaptophysin XTC Restoration. (A) Immunostaining for phosphorylated tau (pTau) and Imaris surface function used to quantify neuritic tau pathology. (B) Synaptophysin immunolabeling with XTC image restoration. Synaptophysin staining is not seen in neuronal cell bodies in the dentate granule cell layer at the bottom of the image. Scale bars are 10 µm.

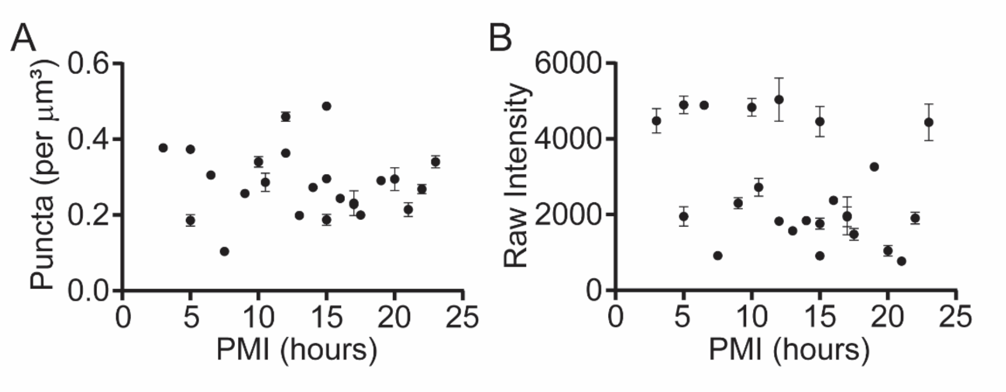


**Supplemental Figure 2**. Synaptophysin Signal Not Associated with Post-Mortem Interval. Post-mortem interval was not associated with either the synaptophysin puncta density ($p$=0.67, simple linear regression) in the dentate gyrus (A) or synaptophysin fluorescence intensity ($p$=0.09, simple linear regression) in the inner molecular layer of the dentate gyrus (B). PMI, post-mortem interval.

**Supplemental Table 1. Imaris Surface Parameters**

| Tau Surface Parameters | Case Settings | Alternate Case Settings[a] |
|---|---|---|
| Enable Region Of Interest | false | false |
| Enable Region Growing | false | false |
| Enable Tracking | false | false |
| Enable Classify | false | false |
| Enable Shortest Distance | true | true |
| Enable Smooth | true | true |
| Surface Grain Size | 0.450 µm | 0.450 µm |
| Enable Eliminate Background | true | true |
| Diameter Of Largest Sphere | 2.00 µm | 1.00 µm |
| Enable Automatic Threshold | false | false |
| Manual Threshold Value | 400 | 450 |
| Active Threshold | true | true |
| Enable Automatic Threshold B | true | true |
| Area above | 4.00 um^2 | 4.20 um^2 |
| Intensity StdDev above | 750 | 750 |
| Intensity Sum above | 6.50e4 | 6.50e4 |

[a]One case required modification of the surface settings to eliminate edge effect which was not present in other cases.
